# Effect of daily consumption of whole fruit feijoa powder on metabolic profile: A Randomised Controlled Trial

**DOI:** 10.64898/2026.09.13.26362958

**Authors:** Aahana Shrestha, Ashish Sarvate, Dilip Mehta, Neeta Nargundhar, Deepak Deshmukh, Chhaya Godse

## Abstract

**Background:** Prediabetes is an intermediate state of increased glucose concentrations above the normal range but below the diagnostic threshold for type 2 diabetes (T2D). Adults with overweight and prediabetes are at increased risk of progression to T2D and metabolic imbalance. Feiolix® -a whole-fruit feijoa powder rich in abscisic acid, polyphenols and dietary fibre which may support metabolic health through multiple, potentially complementary pathways.

**Objective:** To evaluate the effects of low and high-dose Feiolix compared with placebo, on HbA1C, glucagon like peptide (GLP-1) and other metabolic parameters

**Methods:** Adults aged 21–65 years with prediabetes i.e. glycosylated hemoglobulin (HbA1c) 5.7–6.4% and overweight with body mass index (BMI)- 25.0–29.9 kg/m² were randomised to receive Feiolix-LD (300 mg/day) or Feiolix-HD (1,150 mg/day), or placebo (maltodextrin) for 12 weeks. Anthropometric measurements were obtained monthly, and fasting blood samples were collected at baseline and 12 weeks post intervention. Postprandial blood samples were collected at baseline and 12 weeks. Outcomes included HbA1c, GLP-1, fasting plasma glucose, insulin resistance lipid markers, body weight and BMI. Analyses were conducted using ANCOVA model with baseline values as covariates.

**Result:** Of 41 individuals screened, 39 were randomised (13 per group); one participant withdrew from the Feiolix-LD. At week 12, HbA1c decreased by 0.5% in the Feiolix-HD and by 0.4% in the Feiolix-LD, with both reductions significantly greater than placebo (p < 0.05); Compared with placebo, Feiolix supplementation significantly lowered fasting plasma glucose, insulin, HOMA-IR, total cholesterol and LDL-cholesterol. The Feiolix-HD attenuated the increase in triglycerides and reduced body weight and BMI by month 2 compared to placebo. GLP-1 concentrations did not differ between groups. However, no change in GLP-1 was observed between the treatments. No treatment-related adverse events were reported.

**Conclusion:** This study supports the potential of Feiolix supplementation to improve metabolic health, including glycemic and lipid outcomes, with additional benefits for body weight and BMI at the higher dose.

## 1 Introduction

Metabolic syndrome (MetS) is a growing global health concern, estimated to affect 1.54 billion adults worldwide in 2023 (Noubiap et al., 2025). It is defined as a cluster of interrelated cardiometabolic abnormalities, including central adiposity, elevated fasting glucose, elevated triglycerides (TG), reduced high density lipoprotein cholesterol (HDL-C), and elevated blood pressure (Alberti et al., 2009). The presence of three or more of these abnormalities is associated with an increased risk of type 2 diabetes (T2D) and cardiovascular disease (Alberti et al., 2009; Grundy et al., 2005).

Prediabetes is an intermediate state characterised by glucose concentrations above the normal range but below the threshold for T2D (Dhruve et al., 2026). Although prediabetes and MetS are distinct clinical entities, they commonly overlap (Diamantopoulos et al., 2006). Adults with overweight and prediabetes are at increased risk of developing T2D and cardiovascular disease (Huang et al., 2016; Zhou et al., 2002).Annual progression from prediabetes to T2D has been estimated at approximately 3.5–7.0%, compared with around 2% among normoglycemic individuals (Larsen & Torekov, 2017); conversely, reversion to normoglycaemia is associated with a lower subsequent risk of T2D. These individuals therefore represent an important target population for early interventions aimed at reducing cardiometabolic risk and preventing progression to T2D, cardiovascular diseases and other co-morbidities.

Lifestyle modification remains the basis of preventing or delaying progression from prediabetes to T2D. Dietary interventions that improve glycemic control, lipid metabolism, and body weight may therefore provide a practical strategy to reduce cardiometabolic risk. In addition to established healthy dietary patterns, there is growing interest in whole-food ingredients that may complement lifestyle approaches through their naturally occurring bioactive compounds (Castro-Barquero et al., 2020; Godos et al., 2017).

Feijoa (*Acca sellowiana*) is a subtropical fruit native to South America and commercially cultivated in New Zealand. It contains a range of potentially bioactive constituents, including polyphenols, flavonoids, tannins, dietary fibre, and abscisic acid (ABA), which may collectively support glycaemic and lipid regulation. Polyphenols may reduce oxidative stress and low-grade inflammation associated with insulin resistance, while dietary fibre may moderate glucose and lipid absorption and influence gut microbiota metabolism. ABA may further contribute to glucose homeostasis through insulin-independent glucose uptake and incretin-related pathways, including stimulation of glucagon-like peptide-1 (GLP-1) secretion.

Preclinical evidence supports the potential metabolic activity of feijoas. In high-fat diet (HFD) induced diabetic mice, freeze-dried feijoa powder improved fasting glucose, HbA1c, triglycerides, LDL-cholesterol, and total cholesterol, with beneficial effects observed at a human-equivalent dose as low as 300 mg (Rosendale et al., 2026). Clinical evidence is however limited to two human clinical studies. A placebo-controlled trial of feijoa pulp extract in adults with T2D reported improvements in HbA1c, fasting glucose, lipid markers, and blood pressure. The extract used was derived from pulp, which may not have retained the dietary fibre and broader bioactive profile of ABA and polyphenols present in whole-fruit powder. A subsequent double-blind trial of whole-fruit feijoa powder in adults with prediabetic obese individuals- the FERDINAND study, reported improvement in systolic blood pressure but no significant change in fasting glucose however, interpretation was limited by a concurrent dietary intervention (Mohamed et al., 2026)

Therefore, the present study evaluated the effects of Feiolix, a freeze-dried whole-fruit feijoa powder, on glycaemic and lipid outcomes in adults with overweight and prediabetes, representing a population with early metabolic dysfunction and elevated cardiometabolic risk. The study was conducted without concurrent dietary modification to isolate the effects of the whole fruit feijoa powder at low and high doses to examine potential effects on glucose management and to explore plausible mechanisms.

## 2 Materials and Methods

### 2.1 Study Designs

This was a 3-month randomised, double-blind, placebo-controlled parallel study investigating whole feijoa fruit powder (Feiolix^®^) in individuals with prediabetes. The clinical trial was prospectively registered in https://ctri.nic.in under CTRI Registration No: CTRI/2025/08/093439. Written informed consent was obtained from eligible participants prior to study commencement.

Subjects were recruited from the database of the Principal Investigator for the study between 30-Sep-2025 and 27-Oct-2025. A total of 41 participants were screened, and 39 eligible participants were randomised in the ratio 1:1:1 into 3 arms. 1) Feiolix-High dose (1150 mg), 2) Feiolix-Low dose (300 mg) or 3) placebo (maltodextrin) using a randomisation schedule. The randomization schedule was generated using SAS®. Computer-generated block randomization was used to randomize the participants eligible for inclusion for three study groups. Equal allocation of participants in each sequence was ensured. One participant withdrew consent after randomisation. (Figure 1)

**Figure 1.**
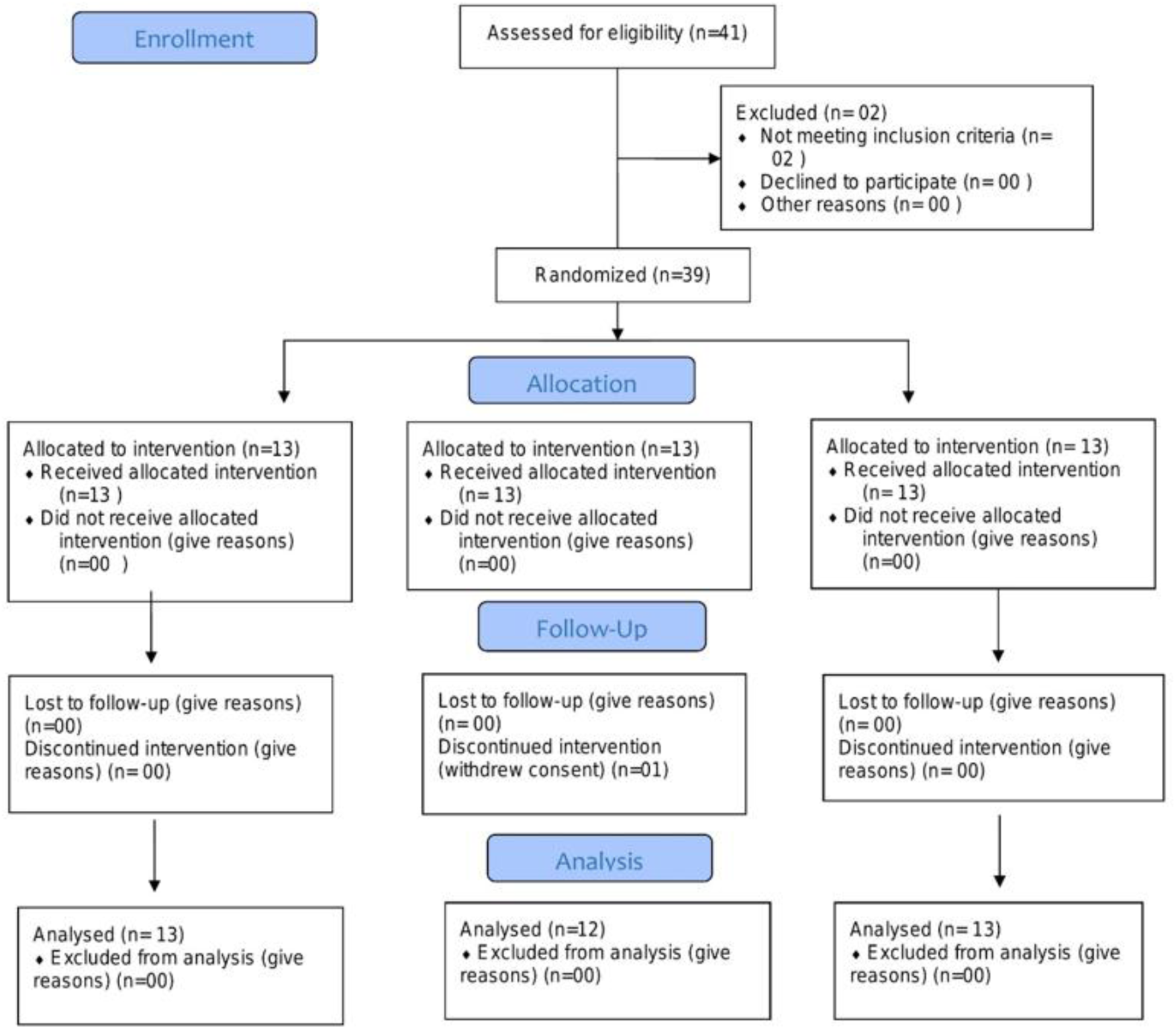
Consort flow diagram describing the recruitment process.

### 2.2 Participants and Procedures

Participants attended the clinic for a screening visit (Visit 1), during which demographic data and medical history were recorded. Laboratory assessments at Visit 1 included glycosylated haemoglobin (HbA1c), complete blood count (CBC), liver function tests (LFT): aspartate aminotransferase (AST) and alanine aminotransferase (ALT), and renal function tests (RFT): serum creatinine and estimated glomerular filtration rate (eGFR), and these measures were repeated at the end-of-study visit (Visit 5; Day 90 ± 3). Thyroid function tests were performed at Visit 1, and a urine pregnancy test was conducted for all women of childbearing potential at screening and again at Visit 5.

Eligible participants were subsequently randomised to receive the investigational product (IP) and attended Visit 2, during which the IP was dispensed. Additional laboratory parameters, including triglycerides, low-density lipoprotein (LDL) cholesterol, high-density lipoprotein (HDL) cholesterol, total cholesterol, insulin, and glucagon were measured at Visit 2 and repeated at Visit 5. Anthropometric measurements (body weight and height) were obtained at Visit 1 and then monthly for three months (Visit 3/Day 30 ± 3, Visit 4/Day 60 ± 3, and Visit 5/Day 90 ± 3). A physical examination was performed at every visit.

At Visit 2 and Visit 5, participants were provided with a standardised breakfast and blood samples were collected at fasting and at 30, 60, and 120-minutes postprandially. Serum glucose and glucagon-like peptide-1 (GLP-1) concentrations were measured at each of these time points. Macro-Nutrient Composition of the Standardised Breakfast is listed in Table 1.

**Table 1.** Composition of standardised breakfast.

| Nutrient | Energy Contribution<br>(% of Total Energy) | Energy (kcal) |
| --- | --- | --- |
| Carbohydrates | 63% | 330.1 |
| Proteins | 19% | 99.6 |
| Fat | 18% | 94.3 |
| Total Energy | 100% | 524 |

#### 2.2.1 Inclusion Criteria

Inclusion criteria for the study comprised the following: individuals with prediabetes (HbA1C level between 5.7% and 6.4 %) and aged between 20-65 years; with a body mass index (BMI) between 25 and 29 kg/m^2^.

#### 2.2.2 Exclusion criteria

The exclusion criteria comprised the following: type 1 or type 2 diabetes mellitus; hypothyroidism or hyperthyroidism; history of bariatric surgery or any other surgery in the last 6 months; use of immunosuppressive medications; history of or ongoing malignancy; abnormal liver or kidney function tests; presence or history of psychiatric disorders, including eating disorders; significant changes in usual diet and/or weight loss of more than 4.5 kg in the last 2 months; known history of chronic smoking, alcohol or drug abuse within the 12 months prior to screening; history of obesity; use of any other phytoherbal, dietary, nutraceuticals, allopathic, or Ayurvedic supplements for weight loss or blood glucose management, or supplements known to affect hunger, satiety, appetite, or the gut microbiome; and pregnancy or lactation.

### 2.3 Interventions

The interventions used were Feiolix- LD, Feiolix- HD or placebo. Feiolix is a freeze-dried whole fruit powder derived from NZ feijoas. Three different interventions were used: 1) Feiolix-LD (one capsule with 300 mg Feiolix powder and three capsules with placebo), 2) Feiolix-HD (four capsules, each containing 287.5 mg Feiolix powder; providing a daily dose of 1150 mg of Feiolix); and 3) placebo (four capsules with each capsules containing 300 mg maltodextrin). The participants were instructed to consume four capsules with water, once a day, 5 minutes before breakfast, throughout the study period. The total duration of treatment was 90 days. The total energy content of the treatments was approximately similar at 1680 kJ per 100 g.

### 2.4 Outcome Measure

The primary outcome was the change in HbA1C after 3 months of Feiolix intervention (Feiolix-HD and Feiolix-LD) compared with placebo. The study also explored postprandial changes in total GLP-1 following the standardised breakfast at baseline and after the Feiolix intervention. Secondary outcomes included fasting serum glucose, insulin, glucagon, ghrelin, HOMA-IR, and lipid profile parameters (Triglycerides (TG), total cholesterol (TC), low density lipoprotein (LDL), high density lipoprotein (HDL). Additionally, anthropometric parameters included body weight (BW), BMI, and waist circumference (WC).

#### 2.4.1 Glycosylated hemoglobulin (HbA1c)

Whole blood was collected in EDTA vacuum tubes (NexPhlebo Diagnostic Pvt. Ltd, Gujarat, India) and HbA1c was analysed using the Lifotronic H8 HbA1c Analyzer, which employs high-performance liquid chromatography (HPLC technology) for the quantitative determination of HbA1c.

#### 2.4.2 Other blood markers

Venous blood was collected in plain vacutainers (NexPhlebo Diagnostic Pvt. Ltd Gujarat, India). Serum was separated by centrifugation (3000 rpm for 15 minutes at 4°C). The separated serum was aliquoted into four aliquotsfor each of the timepoints and stored at −20°C until shipment for biomarker analysis.

Serum glucose was measured using the Agappe Mispa CX4 Clinical Chemistry analyser (Agappe Diagnostics Ltd., Ernakulam, Kerala, India) based on the Glucose Oxidase– Peroxidase (GOD-POD) enzymatic colorimetric method and serum insulin concentrations were measured using the Autolumo A1000 Immunoassay Analyzer, (Autobio Diagnostics Co., Ltd., Zhengzhou, China) employing the Chemiluminescent Microparticle Immunoassay (CMIA) principle. Total GLP-1, and glucagon were measured using commercial sandwich ELISA kits (Lablisa®, LabreCON, India) following the manufacturer’s protocols and calibration standards.

Insulin resistance (HOMA-IR) was calculated using the Homeostasis Model Assessment for Insulin Resistance (HOMA-IR) as follows:

HOMA IR= Fasting insulin (μIU/mL) × Fasting glucose (mg/dL) / 405.

Lipid profile including TC (CHOD-POD method), triglycerides (Glycerokinase Peroxidase method), HDL cholesterol (Direct Method), and LDL cholesterol were analysed using the Agappe Mispa CX4 Clinical Chemistry Analyser. LDL cholesterol was calculated using the Friedewald equation according to the laboratory protocol.

#### 2.4.3 Safety parameters

Venous blood was collected from the participants before the intervention and at Visit 5 (end of 3 months). Blood safety makers, including renal function measures (creatinine and eGFR) and liver function measures (alanine transaminase (ALT), aspartate aminotransferase (AST)), were analysed using the Agappe Mispa CX4 Clinical Chemistry Analyzer based on standard IFCC-recommended kinetic and colorimetric methods. Creatinine was estimated using the sarcosine oxidase method.

### 2.5 Statistical Analysis

The sample size was calculated based on the primary outcome, change in HbA1c. A sample size of 12 participants per group was determined to be required to detect a significant difference in change in HbA1c between treatment and placebo groups, with 80% power and a two-sided α = 0.05. This was based on a standard deviation of 1.32%, derived from a previously published study conducted under comparable conditions (Taghavi et al., 2012)

All statistical analyses were performed using the SAS® System, version 9.1. Statistical analyses were performed using the ANCOVA model, with baseline values included as a covariate, to compare each Feiolix treatment group (Feiolix-LD and Feiolix-HD) with placebo. For the postprandial analysis following the standardised meal, both Treatment and Treatment × Time effects were evaluated, with baseline values included as a covariate. The level of significance was set at p ≤0.05.

#### 2.5.1 Safety Monitoring

Any adverse event during the trial were monitored. Safety measures included LFT: Alanine Aminotransferase (ALT), Aspartate Aminotransferase (AST), RFT (Serum creatinine and e GFR), total blood count, and thyroid function tests. These were measured at baseline and at the end of the intervention.

## 3 Results

### 3.1 Study population

A total of 41 individuals were identified from the study database and screened between 2025 and 2026 (Figure 1). Of these, 39 participants were randomised into three parallel groups: Feiolix low dose (Feiolix-LD; n = 13), Feiolix high dose (Feiolix-HD; n = 13), and placebo (maltodextrin; n = 13). One participant in the Feiolix-LD group withdrew after randomisation; all remaining participants completed the study (Figure 1).

### 3.2 Demographics

Thirty-eight participants aged 41.3 ± 9.5 years completed the study. Baseline sociodemographic and anthropometric characteristics (age, gender, height, weight, and BMI) are presented in Table 2. Analyses confirmed that the groups were similar with no statistically significant differences in all the parameters measured at baseline.

**Table 2.** Baseline characteristics of study participants.

| Baseline Characteristics | Overall<sup>1</sup><br>N=39 | Feiolix-LD<sup>1</sup><br>n=13 | Feiolix-HD<sup>1</sup><br>n=13 | Placebo<sup>1</sup><br>n=13 | P-value <sup>2</sup> |
| --- | --- | --- | --- | --- | --- |
| Age (years)<sup>a</sup> | 41.3(9.5) | 38.5 (8.1) | 45.2 (10) | 40.2 (10) | 0.190 |
| Sex<sup>b</sup> |  |  |  |  | 0.722 |
| Male | 25 (61.5%) | 7 (54%) | 9 (69.2) | 8 (61.5 %) |  |
| Female | 15(38.5 %) | 6 (46 %) | 4 (30.8) | 5 (38.5 %) |  |
| Height (cm)<sup>a</sup> | 163.4 (7.3) | 162.4 (8.5) | 164.2 (7) | 163.5 (6.7) | 0.815 |
| Weight (kg) <sup>a</sup> | 71.9 (5.81) | 72.9 (8.7) | 71.1 (5.2) | 72.7 (3.1) | 0.795 |
| BMI (kg/m2)<sup>a</sup> | 26.9 (1.43) | 27 (1.4) | 26.4 (0.9) | 27.3 (1.7) | 0.207 |

<sup>1</sup> Mean ± SD; n (%)
<sup>a</sup> analysed using one way anova
<sup>b</sup> analysed using chi-square test.

### 3.3 Outcomes measures

#### 3.3.1 Primary outcomes

##### Glycosylated hemoglobulin (HbA1C)

After 3 months of intervention, mean HbA1c decreased with Feiolix-HD (Δ =-0.5%) and Feiolix-LD (Δ =-0.4%), with no change in placebo group. These reductions were statistically significant compared with placebo for both arms (Feiolix-LD vs. placebo: p <0.001; Table 3)

**Table 3.** Change in biochemical markers over 3 months by treatment group.

| | Feiolix-HD | | Within group $\Delta$ | Feiolix-LD | | Within group $\Delta$ | Placebo | | Within group $\Delta$ |
| --- | --- | --- | --- | --- | --- | --- | --- | --- | --- |
|  | Baseline | 3 months |  | Baseline | 3 months |  | Baseline | 3 months |  |
| HbA1C (%) | 5.9 ± 0.1 | 5.4 ± 0.1*** | -0.5 | 6 ± 0.1 | 5.6 ± 0.1*** | -0.4 | 6 ± 0.1 | 6 ± 0.1 | 0 |
| Glucose (mg/dl) | 84.5 ± 2 | 100.3 ± 4.8 | +15.8 | 85.1 ± 1.9 | 92 ± 4.5* | +7 | 81.3 ± 2.1 | 109.4 ± 5.5 | +28 |
| Insulin (μIU/ml)) | 12.8 ± 0.9 | 33 ± 12.2* | +20.2 | 11.3 ± 1 | 17.2 ± 4.9** | +6 | 10.7 ± 0.7 | 85.2 ± 21.3 | +74.5 |
| Homa-IR | 2.7 ± 0.2 | 9.4 ± 4.3* | +6.8 | 2.4 ± 0.2 | 4.2 ± 1.4** | +1.9 | 2.1 ± 0.1 | 24.5 ± 6.7 | +22.4 |
| LDL (mg/dl) | 110.2 ± 5.1 | 100.4 ± 6.8*** | -9.8 | 111.2 ± 9.8 | 95 ± 8.4*** | -16.2 | 102.7 ± 7.3 | 153.4 ± 11.3 | +50.8 |
| TC (mg/dl) | 172.8 ± 5.7 | 176.4 ± 9.6*** | +3.6 | 177.1 ± 9.9 | 167 ± 8.5*** | -10.1 | 166.2 ± 7.8 | 246.5 ± 13.5 | +80.3 |
| TG (mg/dl) | 108.7 ± 4.6 | 145.7 ± 20.8* | +37 | 117.8 ± 4.7 | 152.4 ± 29.9 | +34.6 | 105 ± 6.9 | 222.6 ± 26.6 | +117.6 |
| VLDL (mg/dl) | 21.7 ± 0.9 | 33.7 ± 5.9 | +12 | 23.6 ± 0.9 | 30.5 ± 6 | +6.9 | 21 ± 1.4 | 44.5 ± 5.3 | +23.5 |
| HDL (mg/dl) | 40.9 ± 1.5 | 42.3 ± 2.7 | +1.4 | 42.4 ± 0.8 | 41.6 ± 2.4 | -0.8 | 42.5 ± 0.8 | 48.4 ± 2.4 | +5.9 |
| GLP-1(pg/ml) | 297.3 ± 59.4 | 320.9 ± 45.8 | +23.6 | 300.6 ± 79.5 | 339.2 ± 55.2 | +38.6 | 260.9 ± 29.3 | 360.4 ± 41.7 | +99.6 |
| Glucagon (pg/ml) | 254.4 ± 63.5 | 143.1 ± 18.7 | -111.3 | 193.4 ± 22.7 | 137.5 ± 19.9 | -55.9 | 183.9 ± 24.7 | 129.3 ± 16.4 | -54.6 |

Data presented as means ± SEM. Feiolix-HD group n=13, Feiolix-LD group n=12; Placebo n=13
\*p < 0.05, \*\*p < 0.01, \*\*\*p < 0.001 indicate a statistically significant difference compared with the placebo group; LDL: Low density lipoprotein, VLDL: very low density lipoprotein; TC: Total cholesterol

##### GLP-1

Following the standardised breakfast, GLP-1 levels did not differ between Feiolix intervention (Feiolix-LD or Feiolix-HD) and placebo (Treatment X Time interaction, p=0.563; data not shown), after adjusting for baseline values (Visit 2). Three months of Feiolix intervention did not change fasting GLP-1 levels compared with placebo (Feiolix-HD vs placebo, p=0.315; Feiolix-LD vs placebo p=0.508; Table 3).

#### 3.3.2 Secondary outcomes

##### Glucose, and Homa- IR

Mean fasting serum glucose increased from baseline to 3 months across all intervention groups. The increase was highest in the placebo group (Δ =28 mg/dL), compared with smaller changes in the Feiolix-LD (Δ = +7 mg/dL) and Feiolix-HD (Δ = +15.8 mg/dL) groups. This between-group difference reached statistical significance only for the Feiolix-LD versus placebo comparison (*p* = 0.029), while the Feiolix-HD versus placebo comparison did not reach significance (p >0.05) (Table 3)

Likewise, fasting insulin and insulin resistance (HOMA-IR) increased across all intervention groups; however, the placebo group showed a significantly greater increase in fasting insulin (Δ =+74.5 µIU/mL) and HOMA-IR (Δ =+22.4) compared to Feiolix-HD insulin (Δ = +20.2 µIU/mL) and HOMA-IR (Δ =+6.8 µIU/mL) or Feiolix-LD (Δ = +6 µIU/mL) and HOMA-IR (Δ =+1.9 (p < 0.05; Table 3)

##### Lipid profile

Lipid parameters, including total cholesterol (TC), triglycerides (TG), LDL-cholesterol, and HDL-cholesterol, were assessed at baseline and after 3 months of intervention. Mean LDL-cholesterol decreased in both the Feiolix high-dose (Feiolix-HD; Δ = −9.8 mg/dL) and Feiolix low-dose (Feiolix-LD; Δ = −16.2 mg/dL) groups, whereas it increased in the placebo group (Δ = +50.8 mg/dL). This between-group difference was statistically significant (*p* < 0.001, Table 2).

TG concentrations increased from baseline in all groups; however, the magnitude of increase was significantly reduced with Feiolix supplementation. The increase in the Feiolix-HD group (Δ = +37.0 mg/dL) was significantly smaller than in the placebo group (Δ = +117.6 mg/dL; *p*= 0.04). A comparable, though smaller, reduction was observed in the Feiolix-LD group (Δ = +34.6 mg/dL), which did not reach statistical significance (*p* = 0.061), representing a trend toward a lower TG increase relative to placebo. Mean VLDL-cholesterol and HDL-cholesterol did not differ significantly between either of the Feiolix group and placebo (*p* > 0.05, Table 3).

##### Anthropometric measurement

Body weight and BMI reduced significantly in the Feiolix-HD group at 2 and 3 months (Δ = −1.07 kg and Δ = −0.38 kg respectively) compared with placebo (Δ = −0.75 kg and Δ = −0.27 kg respectively). Although these parameters decreased with Feiolix-LD as well (Δ = −0.98 kg and Δ = −0.35 kg respectively); this did not reach statistical significance (p >0.05; Table 4).

**Table 4.** Changes in anthropometric outcomes over 3 months by treatment group.

| Outcome | Feiolix-HD | | | | | Feiolix-LD | | | | Within-group $\Delta$ | Placebo | | | | |
| --- | --- | --- | --- | --- | --- | --- | --- | --- | --- | --- | --- | --- | --- | --- | --- |
| | Baseline | Visit-3 | Visit-4 | Visit-5 | Within-group $\Delta$ | Baseline | Visit-3 | Visit-4 | Visit-5 | | Baseline | Visit-3 | Visit-4 | Visit-5 | Within-group $\Delta$ |
| BMI (kg/m<sup>2</sup>) | 26.4 ± 0.3 | 26.2 ± 0.3 | 26.1 ± 0.2* | 26.0 ± 0.3 | -0.38 | 27.0 ± 0.4 | 27.1 ± 0.4 | 26.8 ± 0.4 | 26.7 ± 0.3 | -0.35 | 27.3 ± 0.3 | 27.1 ± 0.3 | 27.1 ± 0.5 | 27.0 ± 0.4 | -0.27 |
| Body weight (kg) | 71.1 ± 1.4 | 70.7 ± 1.6 | 70.4 ± 1.5* | 70.1 ± 1.5* | -1.07 | 71.9 ± 2.4 | 71.6 ± 2.4 | 71.3 ± 2.5 | 70.9 ± 0.4 | -0.98 | 72.7 ± 0.1 | 72.4 ± 0.1 | 72.3 ± 0.9 | 72.0 ± 0.9 | -0.75 |
| Waist circumference (cm) | 88.3 ± 1.8 | 87.0 ± 1.8 | 86.3 ± 1.8 | 85.7 ± 1.7 | -2.62 | 93.3 ± 2.6 | 91.5 ± 2.5 | 91.5 ± 2.5 | 90.8 ± 0.3 | -2.54 | 97.1 ± 0.2 | 95.3 ± 0.2 | 94.9 ± 1.9 | 94.4 ± 1.8 | -2.69 |

Data presented as means ± SEM. Feiolix-HD group n=13, Feiolix-LD group n=12; Placebo n=13
\*p < 0.05, \*\*p < 0.01, \*\*\*p < 0.001 indicate a statistically significant difference compared with the placebo group

##### Safety measures

All the safety parameters including LFT, RFT and CBC showed no clinically significant changes before or after the intervention (Table 5).

**Table 5.** Safety measures before and after treatment.

| Analytes | Feiolix-HD |  | Feiolix-LD |  | Placebo |  |
| --- | --- | --- | --- | --- | --- | --- |
|  | Baseline | 3 months | Baseline | 3 months | Baseline | 3 months |
| Haemoglobin (g/dL) | 13.0 ± 1.53 | 12.8 ± 1.69 | 12.8 ± 1.41 | 13.2 ± 1.64 | 12.8 ± 2.21 | 12.8 ± 1.81 |
| RBC (10<sup>6</sup>/μL) | 4.8 ± 0.46 | 4.8 ± 0.47 | 4.9 ± 0.39 | 5.1 ± 0.69 | 4.9 ± 0.50 | 4.7 ± 0.48 |
| WBC (×10<sup>3</sup>/μL) | 6.8 ± 1.16 | 7.2 ± 1.36 | 6.6 ± 1.33 | 6.4 ± 1.26 | 7.5 ± 1.73 | 8.2 ± 2.75 |
| Neutrophils (%) | 55.2 ± 8.55 | 56.6 ± 6.61 | 55.0 ± 8.86 | 52.2 ± 10.25 | 60.3 ± 6.82 | 62.8 ± 7.13 |
| Lymphocytes (%) | 33.6 ± 6.82 | 29.5 ± 4.80 | 31.8 ± 8.31 | 34.0 ± 9.70 | 29.0 ± 7.01 | 26.8 ± 7.46 |
| Monocytes (%) | 6.1 ± 1.78 | 9.0 ± 4.62 | 6.6 ± 2.29 | 8.8 ± 4.25 | 6.6 ± 2.60 | 7.6 ± 2.96 |
| Eosinophils (%) | 5.1 ± 5.37 | 4.9 ± 5.72 | 6.6 ± 5.86 | 5.2 ± 4.53 | 4.1 ± 2.96 | 2.9 ± 2.42 |
| Platelets (×10<sup>3</sup>/μL) | 240.5 ± 46.78 | 267.7 ± 68.23 | 299.5 ± 79.31 | 286.2 ± 72.26 | 317.5 ± 72.79 | 304.8 ± 83.31 |
| AST (U/L) | 19.4 ± 4.15 | 21.8 ± 4.72 | 19.6 ± 6.28 | 22.0 ± 7.57 | 21.3 ± 3.24 | 28.4 ± 10.70 |
| ALT (U/L) | 21.7 ± 9.71 | 22.1 ± 8.09 | 24.9 ± 12.90 | 20.3 ± 6.03 | 28.2 ± 8.15 | 32.6 ± 21.49 |
| Creatinine (mg/dL) | 0.8 ± 0.12 | 0.8 ± 0.10 | 0.7 ± 0.14 | 0.8 ± 0.12 | 0.8 ± 0.16 | 0.9 ± 0.11 |
| eGFR (mL/min/1.73 m<sup>2</sup>) | 107.3 ± 13.77 | 104.0 ± 14.94 | 114.1 ± 15.46 | 109.1 ± 17.60 | 112.3 ± 12.40 | 99.2 ± 17.26 |

Data presented as means ±SEM.

## 4 Discussion

This study showed that three months of Feiolix, a freeze-dried feijoa powder, produced favourable metabolic effects at both the low (300 mg/day) and high (1,150 mg/day) doses. Compared with placebo, both doses improved longer-term glycaemic control, as reflected by HbA1c, and improved lipid markers, including LDL-cholesterol and total cholesterol. Additional improvements in body weight, BMI, and TG were observed only in the high-dose group. Total GLP-1 and glucagon concentrations did not differ between either Feiolix group and placebo.

The findings from this study are broadly consistent with a dose-related responses reported in a high-fat-diet diabetic mouse model, in which feijoa powder at a 300 mg human-equivalent dose (HED) improved fasting glucose, TG, and cholesterol, whereas the 2300 mg HED dose additionally improved body weight and HDL cholesterol (Rosendale et al., 2026). In the present study, the 1150 mg Feiolix dose was associated with significant weight reduction, although HDL cholesterol did not differ from placebo. The lower fasting glucose observed in the low-dose group, but not the high-dose group, should be interpreted cautiously given the substantial day-to-day variability of fasting glucose. In contrast, HbA1c is a more stable measure of longer-term glycaemic exposure (Selvin et al., 2007) and showed favourable effects in both Feiolix groups.

To our knowledge, no previous human clinical studies have evaluated the effects of the whole feijoa fruit. However, a human study with feijoa extract in individuals with diabetes demonstrated significant improvements in HbA1c, fasting glucose, TC, TG, LDL-cholesterol, and systolic blood pressure (Taghavi et al., 2012). Another clinical study, the-FERDINAND study, used freeze dried feijoa powder in a 6-month study comprising 2 months of a low-energy diet [∼840 kJ/day], followed by 4 months of a normal diet. The study showed improvement in systolic blood pressure but no significant changes in the fasting glucose. However, the concurrent low-energy diet with significant change in body weight (∼ 7.2 kg) in the first 2 months (Mohamed et al., 2026)makes it difficult to evaluate contribution of the feijoa powder. Therefore, the present study was designed to evaluate the effect of feijoa powder without concurrent dietary energy restriction, allowing the observed changes to be more directly attributed to the intervention.

Several bioactive components present in Feiolix, including abscisic acid (ABA), polyphenols, and dietary fibres, may plausibly contribute to its observed metabolic effects. This suggests that Feiolix’s benefits could arise from multiple, potentially complementary pathways rather than a single mechanism of action.

ABA occurs naturally in fruits and vegetables and is also produced endogenously by pancreatic β-cells in response to elevated glucose concentrations (Bruzzone et al., 2008). It stimulates insulin-like activity through an insulin-independent mechanism, binding to LANCL2 (lanthionine synthetase C-like 2) receptors to promote glucose uptake independently of insulin signalling (Spinelli et al., 2024).Human and animal studies have further demonstrated beneficial effects of ABA on glycemic control, insulin sensitivity, and lipid metabolism (Leber et al., 2020; Magnone et al., 2018). ABA has also been shown to support glucose homeostasis via the incretin axis, stimulating GLP-1 secretion from intestinal L-cells through a cAMP/PKA-dependent mechanism and enhancing GLP-1 gene transcription, with oral administration increasing circulating GLP-1 and insulin levels in vivo (Bruzzone et al., 2015)

Feijoa fruit contains a diverse polyphenol profile, including ellagitannins, which may contribute to the favourable glycaemic and lipid outcomes observed in this study. Polyphenol-rich interventions have been associated with reduced oxidative stress and inflammation (Hazewindus et al., 2012; Neunert et al., 2015)-key features of metabolic dysfunction. A recent meta-analysis of 281 studies suggests that multiple polyphenols may improve cardiovascular risk markers, including glycaemic and lipid outcomes (Peng et al., 2020). Ellagitannins in feijoa may contribute indirectly to metabolic regulation following their conversion by gut microbiota to urolithins (Kang et al., 2016). Preclinical studies suggest that urolithin can improve insulin sensitivity and mitochondrial function and reduce indices of metabolic inflammation (Toney et al., 2019).Urolithins have also shown anti-inflammatory activity by limiting NF-κB-associated signalling in cellular and animal models (González-Sarrías et al., 2010). Given the established role of chronic NF-κB activation in metabolic inflammation and insulin resistance, this represents one plausible mechanism by which feijoa polyphenols could contribute to the observed metabolic effects. These mechanisms were not directly assessed in the present study and require confirmation in targeted mechanistic studies.

Despite the improvements in glycemic control and insulin resistance observed in the present study, GLP-1 levels did not differ between the Feiolix and placebo groups. This raises two plausible explanations: Feiolix exerts its metabolic effects through alternative pathway (like ABA mediated LANCL2 pathway or polyphenol mediated pathway) rather than the GLP-1: a relevant consideration amid the current focus on GLP-1 receptor-targeted therapies. Alternatively, this finding may reflect methodological limitations rather than a true absence of biological effect. Regarding the latter, the assay used measured total GLP-1 (active and inactive fractions combined), which may have diluted a treatment-specific signal in the active form. In addition, GLP-1’s short circulating half-life means the postprandial sampling timepoint(s) may not have captured peak secretion. Future studies are warranted to understand the exact mechanism of action of Feiolix by incorporating active GLP-1 assays and more frequent postprandial sampling. Regardless of the mechanism of action, Feiolix consistently improved metabolic health markers. Notably, this benefit was achieved without the GLP-1-related side effects (e.g., gastrointestinal symptoms) that often accompany elevated GLP-1 activity, supporting a favourable efficacy-tolerability profile.

Feijoa contains insoluble and soluble fibres, including xyloglucans, which may delay glucose and lipid absorption and promote bile-acid and cholesterol excretion. Consistent with this, Wang et al. found that bread supplemented with non-digestible feijoa fiber showed reduced *in vitro* glucose release and enhanced bile acid and cholesterol adsorption, suggesting potential hypoglycemic and lipid-lowering effects(Wang et al., 2023). Xyloglucans may also support *Bacteroides* growth and short-chain fatty-acid production, including propionate, which may influence hepatic lipid metabolism and cholesterol regulation (Bell et al., 2018).

Notably, previous studies demonstrating positive metabolic effects with isolated polyphenols, ABA, or fiber typically used considerably higher doses than the low dose of feijoa powder used in the present study (300 mg/day), suggesting that the complex whole-fruit matrix may confer a synergistic benefit beyond that of its individual bioactive components.

Despite the robust design and favourable changes observed, several limitations should be noted. First, the study was powered for the primary outcome of HbA1c, which may have been underpowered to detect differences in secondary outcomes, particularly GLP-1 and glucagon. Second, quantifying GLP-1 posed methodological challenges. While active GLP-1 would have been the ideal target, its physiological concentration is extremely low (0–15 pmol/L) and difficult to reliably detect in a standard commercial laboratory setting. Active GLP-1 is also rapidly degraded by the enzyme DPP-4, with a half-life of only 1–2 minutes, requiring DPP-4 inhibitor collection tubes for accurate measurement. Given these challenges, total GLP-1 was measured instead, consistent with the approach recommended by a systematic review (Larsen & Torekov, 2017).Nonetheless, although the precise mechanism underlying Feiolix’s glycemic and lipid benefits could not be definitively established in this study, the overall findings support a beneficial effect of Feiolix on several cardiometabolic risk markers in individuals with prediabetes without any adverse effects.

## 5 Conclusions

Feiolix, a whole-fruit feijoa powder, may support cardiometabolic health in overweight adults with prediabetes. At a relatively low dose of 300 mg/day, Feiolix was associated with improved glycemic control and insulin resistance, and favourable changes in LDL cholesterol and TC compared with placebo. The higher dose of Feiolix was additionally associated with reductions in body weight, BMI, and TG. Collectively, these results suggest that Feiolix may be a natural supplement that can support glycemic and lipid management in individuals at elevated cardiometabolic risk. Larger, longer-term trials are warranted to confirm these benefits and clarify the underlying mechanisms of action.

## 6 Declarations

### Trial registration

The clinical trial was prospectively registered in https://ctri.nic.in under CTRI Registration No: CTRI/2025/08/093439.

### Ethics approval and consent to participate

The study was conducted in accordance with the Declaration of Helsinki, and approved by Ethicare Ethics Committee,

### Consent for publication

Not applicable

### Availability of data and materials

The datasets generated during and/or analysed during the current study are available from the corresponding author on reasonable request.

### Author Contributions

Conceptualization: A.S; D.M.; Methodology, C.G. and A.S.S.; formal analysis, C.G.; and A.S.S; Investigation, A.S.S.; N.N.; and D.D.; writing-original draft preparation, AS; writing-review and editing, A.S.; C.G; A.S.S. All authors have read and agreed to the published version of the manuscript.

## Acknowledgments

The authors would like to acknowledge Viridis BioPharma Pvt. Ltd., Mumbai, Maharashtra, India, for their support and funding.

